# Associations of adolescent menstrual symptoms with school absences and educational attainment: analysis of a prospective cohort study

**DOI:** 10.1101/2024.04.24.24306294

**Authors:** Gemma Sawyer, Abigail Fraser, Deborah A. Lawlor, Gemma C. Sharp, Laura D. Howe

## Abstract

**Background:** Menstrual symptoms may negatively impact pupils’ attendance and educational attainment, but rigorous population-based studies are lacking.

**Methods:** In a prospective cohort study in England, we assessed associations of self-reported heavy or prolonged bleeding and menstrual pain with school absences and attainment, adjusting for potential confounders.

**Findings:** Of 2,698 participants, 36% reported heavy or prolonged bleeding and 56% reported menstrual pain. Heavy or prolonged bleeding was associated with missing 1·7 additional days of school per year (16·58%; 95% CI: 9·43, 24·20), and 48% higher odds of being persistently absent (≥10% absent) (OR 1·48; 95% CI: 1·45, 1·52). Menstrual pain was associated with missing 1·2 additional days of school per year (12·83%; 95% CI: 6·14, 19·95), and 42% higher odds of being persistently absent (OR 1·42; 95% CI: 1·39, 1·46). Heavy or prolonged bleeding was associated with lower scores in compulsory examinations taken at age 16 (−5·7 points; 95% CI: −10·1, −1·2, equating to one lower grade), and with 27% lower odds of achieving five standard passes (OR 0·73; 95% CI: 0·71, 0·75). There was less evidence of an association between menstrual pain and examination scores (−3·14 points; 95% CI: - 7·46, 1·17), but menstrual pain was associated with 16% lower odds of achieving five standard passes (OR 0·84; 95 CI: 0·81, 0·86).

**Interpretation:** Both heavy or prolonged bleeding and menstrual pain were associated with lower school attendance and educational attainment. Greater research and support are needed to enable girls to achieve their full academic potential.

**Funding:** Wellcome Trust and UK Medical Research Council.

## Introduction

Menstrual symptoms, such as pain (dysmenorrhea) and heavy menstrual bleeding (HMB) can affect a high proportion of girls and people who menstruate and have the potential to negatively impact their health, wellbeing, and ability to attend school.^1–7^ A systematic review of cross-sectional studies reported that 12% of school/university students aged 18 and under in high-income countries, and 26% in low- and middle-income countries, have been absent from school due to dysmenorrhea.^8^ Menstrual symptoms could also impair the ability to fully concentrate or participate at school. The systematic review also found that up to 41% of students reported that menstrual pain negatively impacts their performance.^8^ Studies in women of reproductive age have also demonstrated associations between menstrual symptoms and impaired productivity at school or work.^9,10^ Thus, absences and impaired ability to engage with schoolwork could lead to effects of menstrual symptoms on educational attainment.

However, the existing literature on menstrual symptoms and school attendance and attainment is limited. The research primarily consists of cross-sectional surveys recruited specifically with a focus on menstruation where participants simultaneously self-reported their menstrual symptoms and absences/productivity, increasing the likelihood of both selection and misclassification bias.^8^

Examining this relationship longitudinally, within a population sample, where menstrual symptoms have been reported prior to absences/productivity will minimise these issues. It is also possible that students may underreport absences because they either do not feel comfortable disclosing the reason for their absence or may not be aware that their menstrual symptoms have resulted in them feeling unwell (e.g., fatigue from iron deficiency anaemia because of HMB).^6^ It would therefore be useful to examine how menstrual symptoms relate to school-recorded, as opposed to self-reported, absences.

Similarly, previous research has focused on self-reported concentration or productivity.^8^ Whilst it is important to research perceived experiences, it would also be useful to examine the relationship between menstrual symptoms and examination results to understand how experiences of reduced productivity translate into educational attainment. An additional issue is that most research focuses on pain or menstruation-related symptoms broadly. The literature on HMB is comparatively scarce despite HMB contributing to menstrual anxiety, a greater need for access to toilets and menstrual products, and physical and cognitive symptoms related to iron deficiency.^6^ Finally, as few studies have adjusted for key confounders, notably socioeconomic position and early life mental health, it is unclear whether relationships between menstrual symptoms and educational outcomes are causal.

The current study aims to explore the associations between HMB and menstrual pain, and school absences and educational attainment. To achieve this, we use symptom data from a UK longitudinal birth cohort and linked administrative data on school absences and attainment.

## Method

We have followed the STROBE (Strengthening the Reporting of Observational Studies in Epidemiology) guidelines in the reporting of this study.^11^

### Participants

The Avon Longitudinal Study of Parents and Children (ALSPAC) is a longitudinal birth cohort that recruited pregnant women resident in Avon, UK with expected delivery dates between 1st April 1991 and 31st December 1992. The initial number of pregnancies enrolled was 14,541, with 13,988 children alive at age 1. When the oldest children were approximately 7 years of age, an attempt was made to bolster the initial sample with eligible cases who had not joined the study originally. Including children recruited at age 7, the total cohort size is 14,901 children who were alive at 1 year of age.

At age 18, study participants were sent ‘fair processing’ materials describing ALSPAC’s intended use of their administrative records and were given clear means to consent or object via a written form. Administrative data were not extracted for participants who objected, or who were not sent fair processing materials. Ethical approval for the study was obtained from the ALSPAC Ethics and Law Committee and the Local Research Ethics Committees (NHS Haydock REC: 10/H1010/70). Informed consent for the use of data collected via questionnaires and clinics was obtained from participants. Further details on ALSPAC have been published elsewhere.^12,13^ The study website contains details of all the data that is available through a fully searchable data dictionary and variable search tool: http://www.bristol.ac.uk/alspac/researchers/our-data/.

Figure 1 shows the flow of participants from 7,225 offspring with female sex (assigned at birth) who were alive at age one, and hence potentially eligible to be included in this study, to the 2,698 (37%) who had data on all exposures and outcomes.

**Figure 1.**
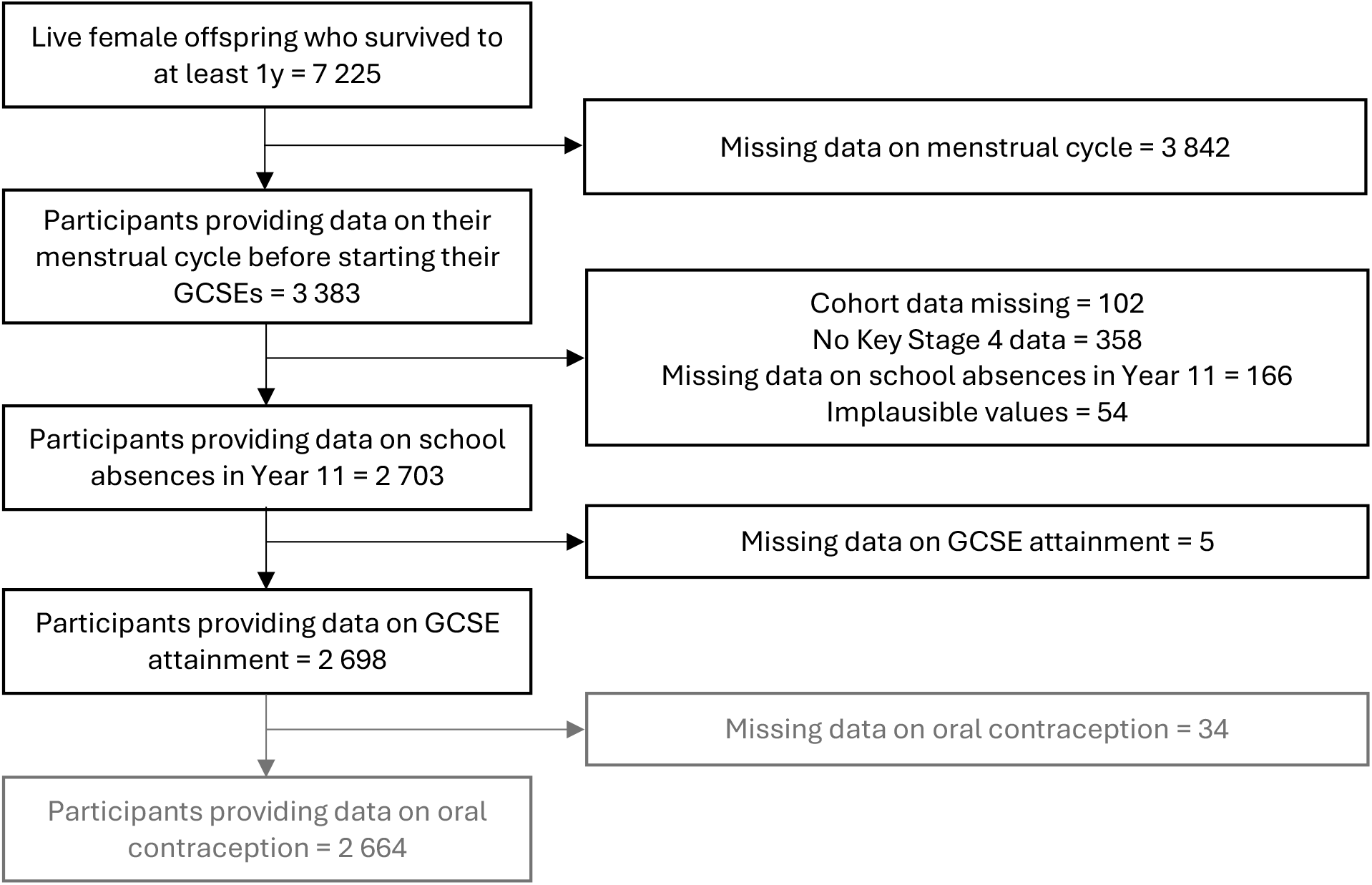
Flow Diagram of the Avon Longitudinal Study of Parents and Children Participants into The Current Study Sample.

### Exposures

Participants were asked about menstruation repeatedly throughout adolescence in nine questionnaires.^14^ We defined menstrual symptoms based on the measure closest in time, but no more than two years, before January of the final compulsory year of schooling (September – July, age 15/16). Therefore, menstrual data was extracted from one of four questionnaires and the participants were aged 13 (N=169), 14 (N=501), 15 (N=996), or 16 (N=1032).

#### Heavy or prolonged bleeding

Participants were asked whether or not they had ever experienced “heavy or prolonged bleeding” with their period. These variables were used to derive a binary heavy or prolonged bleeding variable (‘yes’ or ‘no’).

#### Menstrual-related pain

Depending on which of the four questionnaires participants answered, they were either asked whether they had ever experienced “severe cramps” (yes or no) or “pain with their period” and, if yes, whether this pain was mild, moderate, or severe. We derived a binary variable for menstrual pain; ‘severe cramps or moderate/severe pain with period’ or ‘no severe cramps or mild/no pain with period’.^14^

### Outcomes

Outcome data came from linkage to the National Pupil Database (NPD) (https://find-npd-data.education.gov.uk/).

#### Absences

We used absences in the final year of compulsory schooling when pupils take final exit examinations (age 15/16). Absences were defined as the number of half day sessions missed (whether or not these were authorised, e.g. illness reported by the parents, or unauthorised, e.g. term-time holidays or truancy) divided by the total number of available sessions. Participants with implausible values for the total number of available sessions (including zero or more than 330, N=54) were excluded. We also dichotomised absences based on whether participants were persistently absent (defined as 10% or more), following the definition monitored at a school level in the UK.^15^

#### Educational attainment

Educational attainment was based on GCSE (General Certificate of Secondary Education) qualifications, which are compulsory qualifications in a range of subjects usually taken at age 15/16. At the time GCSEs were completed by this cohort, they were graded A* (highest) to G (grade C reflects a standard pass), and U (unclassified). We used GCSE and equivalent total point score, which is a continuous measure (range 0-540), using up to eight highest GCSE grades, where six points are given for each grade increase (grades and score: A*=58; A=52; B=46; C=40; D=34; E=28; F=22; G=16; U=0). Scores above 464 (eight A* grades) reflect pupils who took Advanced Subsidiary level (AS-level) exams early (advanced qualifications usually taken the year after GCSEs for those who continue in education). We also used a binary educational attainment outcome reflecting whether participants achieved five A*-C GCSEs (standard passes), including Maths and English, which was an important performance indicator for this cohort, and a frequently used criteria for further education.^16^

### Confounders

We selected confounders based on whether they are known to, or could plausibly, cause reporting of exposures and outcomes. The selected confounders were parental occupational social class, maternal education, home ownership, financial difficulties, maternal smoking, ethnicity, age at menarche, body mass index (BMI) at age 13, maternal depression, internalising and externalising symptoms at age 10, intelligence quotient (IQ) at age 8, and adverse childhood experiences. Full details are presented in the supplement.

### Statistical analysis

Analyses were conducted in Stata (version 18·0).^17^ Percentage absences had a positively skewed distribution, so we performed a log-transformation. This means that, when absence is the dependent variable, the regression coefficients represent the percentage difference in time absent between the exposed and unexposed groups. Linear (GCSE score and percentage absence) or logistic (‘persistent absence’ and ‘achieved 5 A*-C GCSEs including English and Maths’) regression models were conducted to estimate the association between the exposures and outcomes, adjusting for confounders.

### Missing data

We did not impute exposures and outcomes due to uncertainty about the ability to predict missing values of menstrual symptoms, and high data availability in outcomes due to the use of linked data. Of the 2,698 participants included in our main analyses (with complete data on all exposures and outcomes; Figure 1), 1,424 (52·8%) had missing data on at least one confounder. We used multiple imputation (MI) to address missing confounder data (Table 1); full details are presented in supplementary material.

**Table 1.**
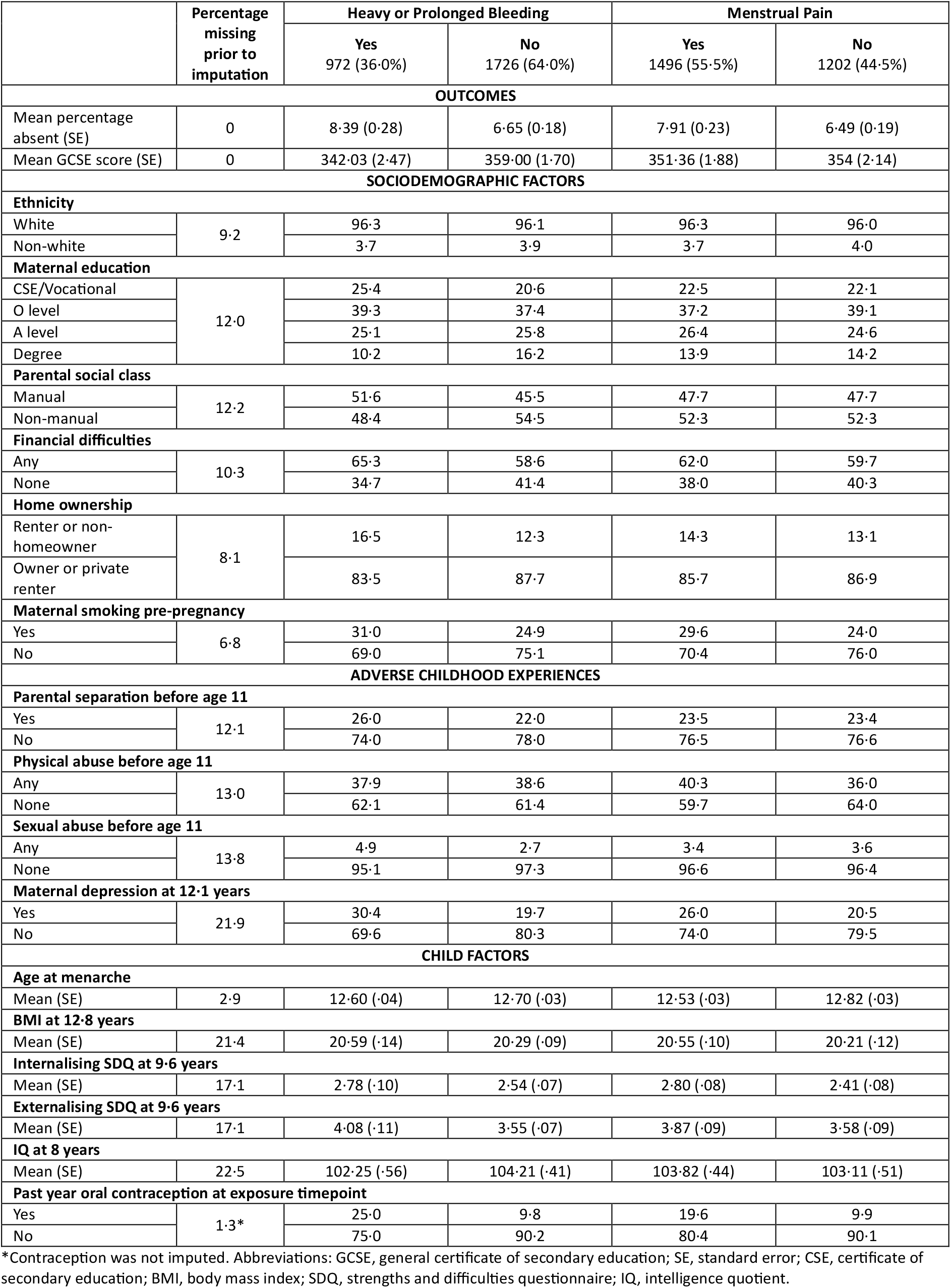
Distribution of outcomes, sociodemographic factors, adverse childhood experiences, and child factors according to adolescent menstrual symptoms.

### Additional analyses

We conducted additional analyses to explore whether our results were influenced by the specific definitions of menstrual symptoms. Specifically, using the imputed data, we conducted confounder adjusted linear regression to assess the associations between four exposures and the continuous outcomes:

a. A three-level variable based on whether participants went to the doctor for heavy or prolonged bleeding (‘heavy or prolonged bleeding and went to the doctor’, ‘heavy or prolonged bleeding but did not go to the doctor’, or ‘no heavy or prolonged bleeding’) to explore whether any effects of heavy or prolonged bleeding are more severe for participants who sought medical help.
b. A three-level variable based on whether participants went to the doctor for menstrual pain (‘pain and went to the doctor’, ‘pain but did not go to the doctor’, or ‘no pain’) to explore whether any effects of menstrual pain are more severe for participants who sought medical help.
c. A four-level variable to separate heavy bleeding from prolonged bleeding. Participants were asked how many days bleeding they usually have during each period and were able to report the exact number of days or, if they were unsure, select one of three categories: 3 days or less, 4-6 days, or 7 days or more. This was used alongside responses to the main heavy or prolonged bleeding variable to derive a four-level variable: ‘heavy and prolonged bleeding’ (‘yes’ to heavy or prolonged bleeding AND reporting 7 days or more bleeding), ‘heavy bleeding only’ (‘yes’ to heavy or prolonged bleeding, BUT reporting 6 or fewer days bleeding), ‘prolonged bleeding only’ (‘no’ to heavy or prolonged bleeding, BUT reporting 7 days or more bleeding), or ‘neither heavy nor prolonged’ (‘no’ to heavy or prolonged bleeding AND reporting 6 days or fewer bleeding’).
d. A four-level variable to explore the effects of co-occurring menstrual symptoms: ‘heavy bleeding and pain’, ‘heavy bleeding only’, ‘pain only’, and ‘neither heavy bleeding nor pain’.

### Oral contraceptive use

Oral contraceptives are often prescribed to ameliorate menstrual symptoms and, could potentially impact absences or attainment indirectly through mood, physical, or cognitive side effects. Although we do not have data on the relative timing of contraceptive use and menstrual symptoms, and thus it is difficult to identify if and how contraception should be accounted for in the analysis, it is more likely that the menstrual symptom preceded contraceptive use as participants are reporting whether they have *ever* experienced menstrual symptoms (compared with contraception use in the last 12 months), meaning adjusting for contraceptive use would be inappropriate over-adjustment.

However, contraceptive use may modify the effect of menstrual symptoms on educational outcomes; i.e., in people who report ‘ever’ experiencing the symptom but whose symptoms have improved following contraception initiation, educational consequences may be diminished. Therefore, in a final sensitivity analysis, we stratified the analyses by past year oral contraceptive use (‘oral contraceptive pill’ or ‘no oral contraceptive pill’), which was self-reported in the same questionnaire as the participants reported their menstrual symptoms. This was conducted in participants with complete data on contraceptive use only (Figure 1), using a likelihood ratio test to assess statistical evidence of interaction.

## Results

Of the 2,698 participants, 972 (36%) reported heavy or prolonged bleeding and 1,496 (55%) reported menstrual pain (severe cramps or moderate/severe pain with their period). Table 1 shows the proportion or mean (standard deviation) of outcomes and covariates for the groups with and without each menstrual symptom. The participants included in the analyses tended to be of higher socioeconomic position than those excluded due to missing data (Supplementary Table 2). Figure 2 shows results of the unadjusted and adjusted regression models. All results (imputed and complete case) are presented in Supplementary Tables 3-7.

**Table 2.** Linear regression analysis of the associations between menstrual symptoms and school absences and GCSE score stratified by past year oral contraceptive use in the sample providing data on oral contraceptive use (N=2664).

|  | School absence in Year 11 |  |  |  | GCSE points score |  |  |  |
| --- | --- | --- | --- | --- | --- | --- | --- | --- |
|  | % change | 95% CI | P value | Interaction p value | Beta | 95% CI | P value | Interaction p value |
| HEAVY OR PROLONGED BLEEDING |  |  |  |  |  |  |  |  |
| Crude Analysis (N=2664) |  |  |  |  |  |  |  |  |
| Data on oral contraceptive use (N=2664) | 22.41 | 14.87, 30.44 | <0.0001 | 0.8816 | -17.36 | -23.12, -11.60 | <0.0001 | 0.7561 |
| No oral contraceptive use (N=2257) | 16.11 | 8.26, 24.52 | <0.0001 |  | -14.59 | -20.83, -8.35 | <0.0001 |  |
| Oral contraceptive use (N=407) | 17.63 | 0.03, 38.33 | 0.0495 |  | -12.11 | -28.44, 4.22 | 0.1457 |  |
| Adjusted Analysis (N=1258) |  |  |  |  |  |  |  |  |
| Data on oral contraceptive use (N=1258) | 11.27 | 1.74, 21.70 | 0.0195 | 0.7104 | -6.31 | -11.83, -0.79 | 0.0250 | 0.6601 |
| No oral contraceptive use (N=1093) | 7.79 | -2.22, 18.84 | 0.1314 |  | -5.21 | -11.07, 0.65 | 0.0813 |  |
| Oral contraceptive use (N=165) | 14.92 | -8.93, 45.02 | 0.2394 |  | -2.55 | -19.00, 13.90 | 0.7600 |  |
| MENSTRUAL PAIN |  |  |  |  |  |  |  |  |
| Crude Analysis (N=2664) |  |  |  |  |  |  |  |  |
| Data on oral contraceptive use (N=2664) | 16.33 | 9.38, 23.71 | <0.0001 | 0.5506 | -3.10 | -8.69, 2.50 | 0.2784 | 0.8483 |
| No oral contraceptive use (N=2257) | 12.24 | 5.14, 19.82 | 0.0005 |  | -0.24 | -6.09, 5.61 | 0.9351 |  |
| Oral contraceptive use (N=407) | 13.44 | -4.93, 35.37 | 0.1613 |  | -3.75 | -21.55, 14.06 | 0.6795 |  |
| Adjusted Analysis (N=1258) |  |  |  |  |  |  |  |  |
| Data on oral contraceptive use (N=1258) | 11.47 | 2.41, 21.34 | 0.0121 | 0.2510 | -2.61 | -7.84, 2.62 | 0.3284 | 0.2960 |
| No oral contraceptive use (N=1093) | 12.03 | 2.45, 22.50 | 0.0128 |  | -2.35 | -7.74, 3.03 | 0.3915 |  |
| Oral contraceptive use (N=165) | -1.97 | -24.29, 26.91 | 0.8790 |  | 2.59 | -15.58, 20.77 | 0.7783 |  |

Abbreviations: GCSE, general certificate of secondary education. Adjusted for ethnicity; maternal education, parental social class, financial difficulties, and home ownership during pregnancy; maternal smoking pre-pregnancy; parental separation, physical abuse, and sexual abuse before 11; maternal depression at 12.1; age at menarche; body mass index at 12.8; internalising and externalising problems at 9.6; and intelligence quotient at 8.

**Figure 2.**
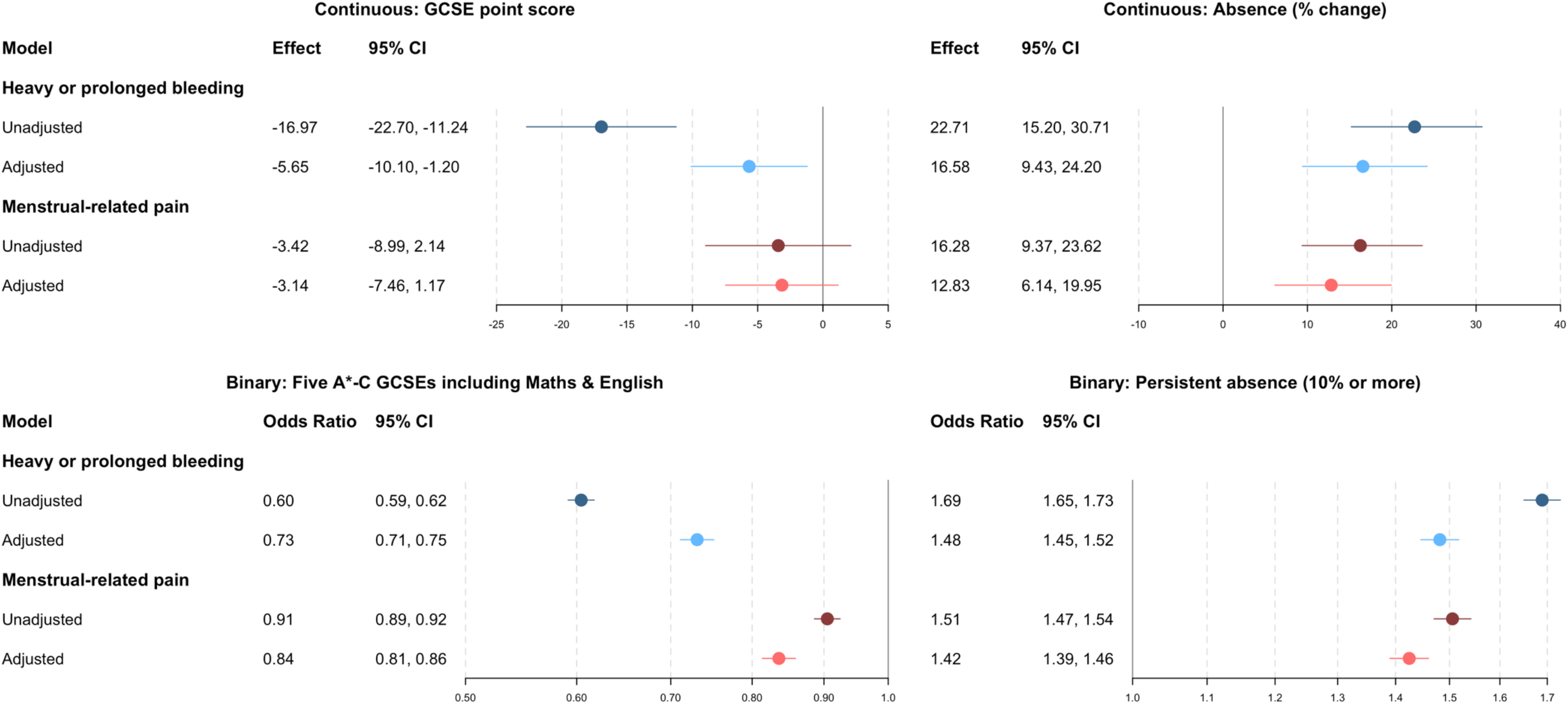
Linear regression analysis of the association between menstrual symptoms and GCSE score and school absences and logistic regression analysis of the association between menstrual symptoms and achieving five A*-C GCSEs including Maths and English and persistent absence (10% or more) (N=2698). Abbreviations: GCSE, general certificate of secondary education. Adjusted for ethnicity; maternal education, parental social class, financial difficulties, and home ownership during pregnancy; maternal smoking pre-pregnancy; parental separation, physical abuse, and sexual abuse before 11; maternal depression at 12·1; age at menarche; body mass index at 12·8; internalising and externalising problems at 9·6; and intelligence quotient at 8.

### Menstrual Symptoms and School Absences

Participants reporting heavy or prolonged bleeding had, on average, 16·6% (95% CI: 9·4, 24·2) more time off school (equating to an average of 1·7 days more) compared to those who had never experienced heavy or prolonged bleeding, after adjusting for all confounders. Participants reporting severe menstrual pain had, on average, 12·8% (95% CI: 9·4, 23·6) more time absent from school (an average of 1·2 days more). When exploring persistent absence (i.e., 10% or more), heavy or prolonged bleeding (OR 1·48; 95% CI: 1·45, 1·52) and menstrual pain (OR 1·42; 95% CI: 1·39, 1·46) were associated with an increased likelihood of being persistently absent (Figure 2). The results from the unadjusted analyses supported the same conclusions.

### Menstrual Symptoms and Educational Attainment

Participants reporting heavy or prolonged bleeding had a lower total GCSE score (−5·65; 95% CI: - 10·10, −1·20) compared with those who did not report heavy or prolonged bleeding, after adjusting for confounders (equivalent to around a 1 grade reduction, i.e., moving from an A to a B). There was less evidence that menstrual pain (−3·14; 95% CI: −7·46, 1·17) was associated with a difference in GCSE score. There was evidence that heavy or prolonged bleeding (OR 0·73; 95% CI: 0·71, 0·75) and menstrual pain (OR 0·84; 95 CI: 0·81, 0·86) were associated with a reduced likelihood of achieving 5 A*-C GCSEs including Maths and English (Figure 2). The results from the unadjusted analyses support the same conclusions.

### Additional Analyses

#### a) Seeking medical care for heavy or prolonged bleeding

The association between heavy and prolonged bleeding and school absences was stronger for those who had seen a doctor about their symptoms (42·6%, 95% CI: 26·4, 60·9; equivalent to 4·1 days) compared with those who reported heavy and prolonged bleeding but had not consulted a doctor (10·9%, 95% CI: 3·7, 18·7; equivalent to 1·0 day). In contrast, the association between heavy and prolonged bleeding with educational attainment was similar whether or not people who had consulted a doctor (−5·5; 95% CI: −10·2, −0·7 versus −5·9; 95% CI: −14·6, 2·7) (Figure 3).

**Figure 3.**
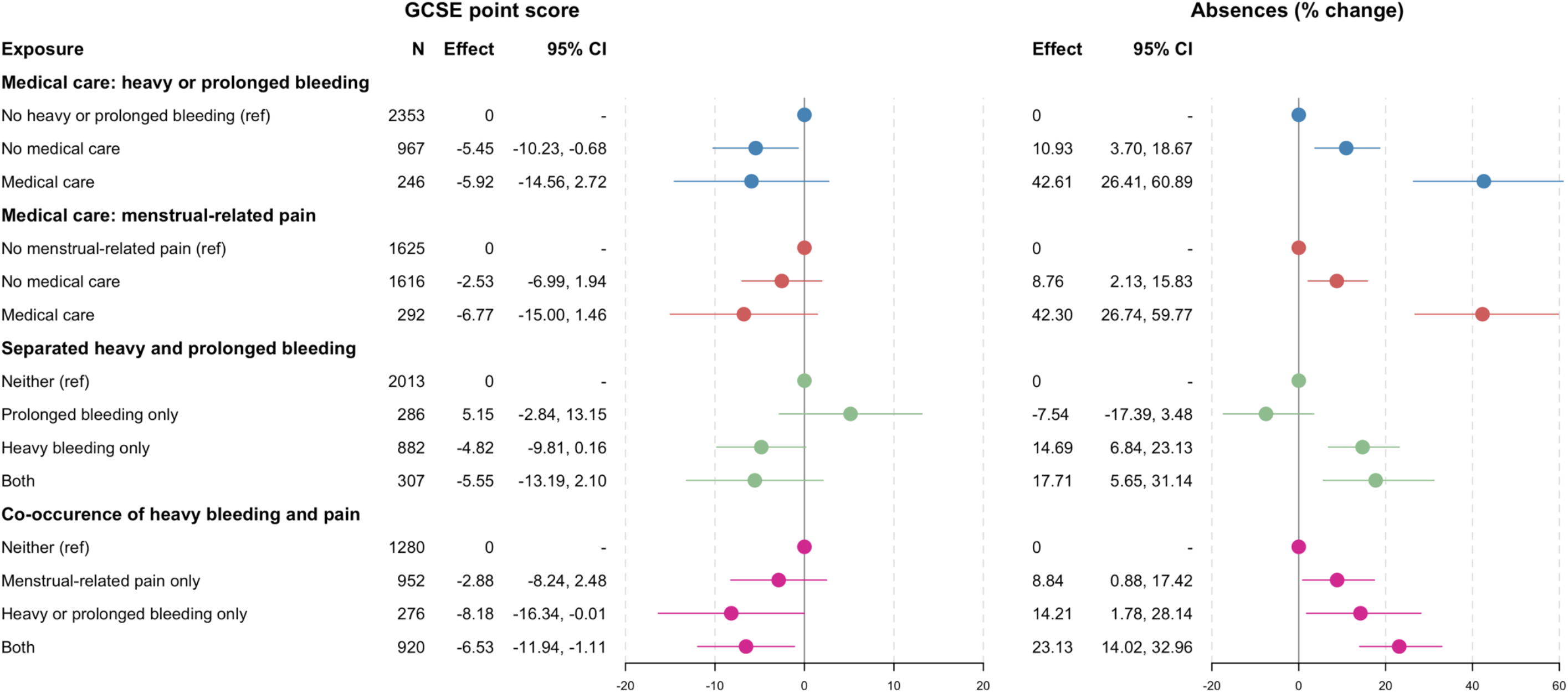
Linear regression analysis of the association between different categories of menstrual symptoms and GCSE score and school absences(N=2698). Abbreviations: GCSE, general certificate of secondary education. Adjusted for ethnicity; maternal education, parental social class, financial difficulties, and home ownership during pregnancy; maternal smoking pre-pregnancy; parental separation, physical abuse, and sexual abuse before 11; maternal depression at 12·1; age at menarche; body mass index at 12·8; internalising and externalising problems at 9·6; and intelligencequotient at 8.

#### b) Seeking medical care for menstrual pain

The association of menstrual pain with absence was stronger in those who had consulted a doctor compared with those who had not consulted a doctor (42·3%; 95% CI: 26·7, 59·8, equivalent to 3·7 days versus 8·8%; 95% CI: 2·1, 15·8, equivalent to 0·8 days). There was weak evidence that pain was associated with educational attainment regardless of doctor consultation, although the effect estimates were slightly larger for those that had compared with had not consulted a doctor (−6·8; 95% CI: −15·0, 1·5 versus −2·5; 95% CI: −7·0, 1·9) (Figure 3).

#### c) Separated heavy bleeding and prolonged bleeding

Participants with ‘heavy bleeding only’ (14·7%; 95% CI: 6·8, 23·1, equivalent to 1·5 days) and ‘heavy and prolonged bleeding’ (17·7%; 95% CI: 5·7, 31·1, equivalent to 1·9 days) were absent from school more than participants who had neither symptom. There was little evidence that participants with ‘prolonged bleeding only’ (−7·5%; 95% CI: −17·4, 3·5) were absent more or less than those with neither symptom. We found minimal evidence that the GCSE score of participants with ‘prolonged bleeding only’ (5·2; 95% CI: −2·8, 13·5), ‘heavy bleeding only’ (−4·8; 95% CI: −9·8, 0·2), or ‘heavy and prolonged bleeding’ (−5·6; 95% CI: −13·2, 2·1) differed compared with those with neither symptom (Figure 3).

#### d) Co-occurrence of heavy or prolonged bleeding and pain

The results suggested that, compared with participants with neither heavy nor prolonged bleeding nor pain, participants in the ‘pain only’ group were absent 8·8% (95% CI: 0·9, 17·4, equivalent to 0·8 days) more, the ‘heavy or prolonged bleeding only’ group 14·2% (95% CI: 1·8, 28·1, equivalent to 1·3 days) more, and the ‘heavy or prolonged bleeding and pain’ group 23·1% (95% CI: 14·0, 33·0, equivalent to 2·2 days) more. There was also evidence that participants in the ‘heavy or prolonged bleeding only’(−8·2; 95% CI: −16·3, 0·0) and ‘heavy or prolonged bleeding and pain’ (−6·5; 95% CI: - 11·9, −1·1) groups had a lower GCSE score than those with neither symptom, but there was little evidence that those in the ‘pain only’ (−2·9; 95% CI: −8·2, 2·5) group had a different GCSE score compared to those with neither symptom (Figure 3).

### Oral contraceptive use

When stratifying by past-year oral contraceptive use, menstrual pain was associated with higher rates of absence in participants not using contraception (N=1093; 12·0%; 95% CI: 2·5, 22·5), but not in those using contraception (N=165; −2·0%; 95% CI: −24·3, 26·9). For all other exposure-outcome pairs, results were similar in participants who were or were not using oral contraception (Table 2).

## Discussion

This study provides evidence that heavy or prolonged bleeding and menstrual pain in adolescence are associated with greater time absent from school and lower educational attainment.

Our findings are consistent with previous research that reported a relationship between heavy bleeding and school/work performance, ^9^ and between menstrual pain and school absenteeism^8,18^ and productivity. ^8,10^ While we found evidence of an association between menstrual pain and attaining five or more good GCSEs, we did not find strong evidence for an association between menstrual pain and GCSE point score (as a continuum). The reasons for this discrepancy are unclear.

This study’s strengths include the longitudinal population-based design, use of objective outcome measures, comprehensive adjustment for confounders, and multiple analyses exploring whether our definitions of menstrual symptoms influenced our main results. Moreover, as in the majority of research in this field, menstrual symptoms were self-reported, which is increasingly recognised as the most appropriate measure.^19,20^

Participants were asked whether they experienced ‘heavy or prolonged bleeding’; this combined question limits our ability to determine which of these two, separate clinical symptoms drive the observed associations.^20^ When attempting to address this by separating heavy from prolonged bleeding, the results suggests that heavy but not prolonged bleeding was associated with more time absent. Results of this sensitivity analysis were less clear for educational attainment, potentially due to lower statistical power in this analysis.

We had to define ‘prolonged bleeding’ as 7 days or more in this analysis, because this was the highest response category provided to participants, even though prolonged bleeding is defined as 9 days or more clinically.^20^ The main menstrual symptom definitions in this study were also somewhat limited as they were binary, meaning the groups are likely to be heterogenous in terms of the severity of their symptoms and there may be some misclassification. We attempted to address this in sensitivity analyses, which generally showed that educational consequences were more severe for people who consulted a doctor about their symptoms. However, health seeking behaviour may be reflective of other factors such as cultural attitudes, menstrual health knowledge, and socioeconomic position.^21,22^ We also conducted a sensitivity analysis to explore the co-occurrence of heavy or prolonged bleeding and pain, which suggested that both heavy or prolonged bleeding and pain (and their co-occurrence) were associated with absences, but pain only was not associated with GCSE score, consistent with the main findings.

An additional limitation is that participants were asked whether they had *ever* experienced the menstrual symptoms, meaning some participants may no longer be experiencing the symptom at the time of outcome assessment. Furthermore, whilst we attempted to minimise loss to follow up and missing data by using linked education data and multiple imputation of confounders to maintain a larger and more representative sample, only 37% of the eligible sample was included in the analysis.

We also note that the current findings are based upon participants who were taking their GCSEs between 2006 and 2009. Surveys of UK school girls conducted between 2019 and 2023 suggest that menstrual difficulties continue to negatively impact attendance and performance.^18,23,24^ We presented our findings to a patient and public involvement group of adolescent girls currently in secondary school, who reported difficulties focusing during menstruation, restricted toilet access, stigma, and insufficient period product provision. Therefore, it is likely that our findings remain highly relevant to current UK students.

Future research identifying the mechanisms underlying the observed associations and potential solutions is warranted. It is likely that there are a number of mechanisms, including menstrual anxiety and concerns about leaking, feelings of shame and embarrassment due to menstrual stigma, bullying, challenges managing symptoms whilst in school, and difficulties accessing toilets during lessons, as well as the experience of debilitating symptoms.^18,24,25^ Exploring these and other potential mechanisms will aid in the development of school-, family-, and community-based interventions.

In summary, our study suggests that heavy or prolonged bleeding and menstrual pain, are associated with lower school attendance and educational attainment. More research is needed to understand the mechanisms behind these associations, and to develop strategies to tackle menstruation-related inequalities to mitigate negative impacts of menstrual symptoms on education.

## Data sharing

ALSPAC data access is through a system of managed open access. The steps below highlight how to apply for access to the data included in this paper and all other ALSPAC data.

1. Please read the <u>ALSPAC access policy (PDF, 891kB)</u> which describes the process of accessing the data and samples in detail, and outlines the costs associated with doing so.
2. You may also find it useful to browse the fully searchable <u>research proposals database</u>, which lists all research projects that have been approved since April 2011.
3. Please submit your research proposal for consideration by the ALSPAC Executive Committee. You will receive a response within 10 working days to advise you whether your proposal has been approved.
4. If you have any questions about accessing data or samples, please (data) or (samples).

Analysis code is available on GitHub at https://github.com/GemmaS17.

## Data Availability

ALSPAC data access is through a system of managed open access. To find out more and apply for access to the data included in this paper and all other ALSPAC data, please visit the ALSPAC website or.

## Acknowledgements

We are extremely grateful to all the families who took part in this study, the midwives for their help in recruiting them, and the whole ALSPAC team, which includes interviewers, computer and laboratory technicians, clerical workers, research scientists, volunteers, managers, receptionists, and nurses.

