## Supplemental Material for "Associations of adolescent menstrual symptoms with school absences and educational attainment: analysis of a prospective cohort study"

### Supplementary Material

#### Supplementary Methods

##### Definition of confounding variables

We included multiple socioeconomic factors reported by mothers during pregnancy, including occupation (dichotomised into 'manual' or 'non-manual' based on the 1991 British Office of Population and Census Statistics classification), maternal education ('Certificate of Secondary Education/vocational', 'O level', 'A level', or 'degree'), home ownership ('owner or private renter' or 'renter or non-homeowner'), financial difficulties score (dichotomised to 'any difficulties (score of 1 or above)' or 'no difficulties (score of 0)), and smoking pre-pregnancy ('any smoking' or 'no smoking') as confounders. Other confounders include child ethnicity ('white' or 'non-white'), age at menarche, body mass index (BMI; $kg/m^2$) measured at a clinic visit at 12.8 years, maternal-reported depression in the past 2 years ('yes' or 'no') collected when the participant was aged 12.1 years, internalising and externalising problems at 9.6 years (each with a score ranging from 0 to 20 and measured with the Strengths and Difficulties Questionnaire),[1] and intelligence quotient at age 8 (measured with the Weschler Intelligence Scale for Children).[2] Adverse childhood experiences (ACEs), including parental separation, sexual abuse, and physical abuse before age 11, were either reported prospectively by their mother or retrospectively self-reported in adulthood. Multiple questionnaire items corresponded to each ACE (see Supplementary Table 1 for details). Binary variables for each ACE construct ('any' or 'none') were derived for participants who responded to at least 50% of the relevant questions for a given ACE; for participants with fewer than 50% of the relevant questions the ACE was coded as missing.[3]

##### Details of multiple imputation

We included all observed variables used in analyses, including exposures and outcomes, in imputation equations. Additional auxiliary variables, including maternal depression during pregnancy (measured with the Edinburgh Postnatal Depression Scale),[4] BMI at age 7, and SDQ at age 11, were used to impute maternal depression, BMI, and internalising and externalising problems, respectively. Results were calculated across 60 imputed datasets, guided by results of mcerror tests, which assess statistical reproducibility of the imputation,[5] and associations were generated by pooling across these datasets using Rubin's rules.[6] The prevalence/means of each confounder before and after multiple imputation are presented in Supplementary Table

**Supplementary Table 1. Variables used to derive adverse childhood experiences (ACEs), including parental separation, physical abuse, and sexual abuse experienced up to age 11.**

| Variable ID | ACE | Description | Reported | Retrospective | Age reported | Start time period | End time period |
|---|---|---|---|---|---|---|---|
| f228 | Parental separation | Divorce >CH born | Parent | No | 8m | 0 | 0.67 |
| f237 | Parental separation | Separation from PTNR >CH born | Parent | No | 8m | 0 | 0.67 |
| g308 | Parental separation | Mum divorced >CH8MTHs | Parent | No | 1yrs9m | 0.67 | 2 |
| g317 | Parental separation | Mum and partner separated >CH8MTHs | Parent | No | 1yrs9m | 0.67 | 2 |
| h218 | Parental separation | Whether mum got divorced since study child was 18 months old and effect this had | Parent | No | 2yrs9m | 1.5 | 3 |
| h227 | Parental separation | Whether mum and partner separated since study child was 18 months old and effect this had | Parent | No | 2yrs9m | 1.5 | 3 |
| j308 | Parental separation | MUM Divorced> CH 30 MTHs | Parent | No | 3yrs11m | 2.5 | 4 |
| j317 | Parental separation | MUM & PTR Separated> CH 30 MTHs | Parent | No | 3yrs11m | 2.5 | 4 |
| k4008 | Parental separation | Mother was divorced in past year | Parent | No | 5yrs1m | 4 | 5 |
| k4017 | Parental separation | Mother and partner separated in past year | Parent | No | 5yrs1m | 4 | 5 |
| l4008 | Parental separation | Respondent was divorced since study child's 5th birthday | Parent | No | 6yrs1m | 5 | 6 |
| l4017 | Parental separation | Respondent separated from partner since study child's 5th birthday | Parent | No | 6yrs1m | 5 | 6 |
| p2008 | Parental separation | Mother was divorced since the study child's 6th birthday | Parent | Yes | 9yrs2m | 6 | 7 |
| p2017 | Parental separation | Mother and husband/partner separated since the study child's 6th birthday | Parent | Yes | 9yrs2m | 6 | 7 |
| r5008 | Parental separation | Respondent has been divorced since child's 9th birthday | Parent | Yes | 11yrs2m | 9 | 10 |
| r5017 | Parental separation | Respondent separated from husband/partner since the study child's 9th birthday | Parent | Yes | 11yrs2m | 9 | 10 |
| f246 | Physical abuse | PTNR physically cruel to CHDR >CH born | Parent | No | 8m | 0 | 0.67 |
| f247 | Physical abuse | MUM physically cruel to CHDR >CH born | Parent | No | 8m | 0 | 0.67 |
| g326 | Physical abuse | Partner physically cruel to children >CH8MTHs | Parent | No | 1yrs9m | 0.67 | 2 |
| g327 | Physical abuse | Mum physically cruel to children >CH8MTHs | Parent | No | 1yrs9m | 0.67 | 2 |
| h236 | Physical abuse | Whether partner was physically cruel to children since study child was 18 months old and effect this had | Parent | No | 2yrs9m | 1.5 | 3 |
| h237 | Physical abuse | Whether mum was physically cruel to children since study child was 18 months old and effect this had | Parent | No | 2yrs9m | 1.5 | 3 |
| j326 | Physical abuse | PTR PHYS Cruel to CDRN> CH 30 MTHs | Parent | No | 3yrs11m | 2.5 | 4 |
| j327 | Physical abuse | MUM PHYS Cruel to CDRN> CH 30 MTHs | Parent | No | 3yrs11m | 2.5 | 4 |
| k4026 | Physical abuse | Mothers partner was physically cruel to children in past year | Parent | No | 5yrs1m | 4 | 5 |
| k4027 | Physical abuse | Mother was physically cruel to children in past year | Parent | No | 5yrs1m | 4 | 5 |
| l4026 | Physical abuse | Respondent's partner physically cruel to respondent's children since study child's 5th birthday | Parent | No | 6yrs1m | 5 | 6 |
| l4027 | Physical abuse | Respondent physically cruel to own children since study child's 5th birthday | Parent | No | 6yrs1m | 5 | 6 |
| p2026 | Physical abuse | Mother's husband/partner was physically cruel to her children since the study child's 6th birthday | Parent | Yes | 9yrs2m | 6 | 7 |
| p2027 | Physical abuse | Mother was physically cruel to her children since the study child's 6th birthday | Parent | Yes | 9yrs2m | 6 | 7 |
| r5026 | Physical abuse | Respondent's husband/partner was physically cruel to their children since study child's 9th birthday | Parent | Yes | 11yrs2m | 9 | 10 |
| r5027 | Physical abuse | Respondent was physically cruel to their children since the study child's 9th birthday | Parent | Yes | 11yrs2m | 9 | 10 |
| ypb8002 | Physical abuse | Frequency adult in family pushed, grabbed or shoved respondent before age of 11 | Child | Yes | 22yrs | 0 | 11 |
| ypb8003 | Physical abuse | Frequency adult in family smacked respondent for discipline before age of 11 | Child | Yes | 22yrs | 0 | 11 |
| ypb8007 | Physical abuse | Frequency adult in family hit respondent so hard it left bruises or marks before age of 11 | Child | Yes | 22yrs | 0 | 11 |
| kf455a | Sexual abuse | Child sexually abused > 18 months, Y/N | Parent | No | 30m | 1.5 | 2.5 |
| kj465 | Sexual abuse | CH was Sexually Abused Past 12 MTHs | Parent | No | 42m | 2.5 | 3.5 |
| kl475 | Sexual abuse | Child was sexually abused since age 3 | Parent | No | 57m | 3 | 5 |

| | | | | | | | |
|---|---|---|---|---|---|---|---|
| kn4005 | Sexual abuse | Child sexually abused in past 15 months | Parent | No | 69m | 4.5 | 6 |
| kq365 | Sexual abuse | Child was sexually abused since his/her 5th birthday | Parent | No | 81m | 5 | 7 |
| kt5005 | Sexual abuse | Since 7th birthday child has been sexually abused | Parent | No | 105m | 7 | 9 |
| ypb8030 | Sexual abuse | Respondent was touched in a sexual way by adult or older child, or was forced to touch adult or older child in a sexual way, before age of 11 | Child | Yes | 22yrs | 0 | 11 |
| ypb8040 | Sexual abuse | Adult or older child forced, or attempted to force, respondent into any sexual activity by threatening or holding respondent down or hurting respondent in some way, before age of 11 | Child | Yes | 22yrs | 0 | 11 |

Information provided by Houtepen et al. (2018).[3]

**Supplementary Table 2. Proportion of missing data in the sample of individuals who provided data on exposures and outcomes (N= 2698) and comparison of prevalence of confounders in the incomplete and imputed samples, and the full female cohort.**

| Variable | Prevalence/mean (SD) in sample restricted to those reporting on exposures and outcomes | Proportion missing data in sample restricted to those reporting on exposures and outcomes | Prevalence/mean in imputed dataset | Prevalence/mean (SD) in the full female cohort |
|---|---|---|---|---|
| Non-white ethnicity | 3.72% | 9.23% | 3.87% | 5.05% |
| Maternal education | 21.10% CSE<br>38.16% O level<br>26.16% A level<br>14.57% Degree | 12.01% | 22.37% CSE<br>38.09% O level<br>25.51% A level<br>14.02% Degree | 25.14% CSE<br>36.89% O level<br>24.01% A level<br>13.96% Degree |
| Manual parental social class | 46.58% | 12.16% | 47.76% | 49.23% |
| Any financial difficulties | 60.55% | 10.27% | 61.06% | 63.92% |
| Non-homeowner or renter | 13.55% | 8.08% | 13.79% | 19.90% |
| Any maternal smoking pre-pregnancy | 26.93% | 6.82% | 27.11% | 33.02% |
| Any maternal depression | 22.78% | 21.91% | 23.62% | 23.29% |
| Any parental separation | 22.82% | 12.12% | 23.53% | 24.95% |
| Any sexual abuse | 3.31% | 13.83% | 3.53% | 3.15% |
| Any physical abuse | 38.76% | 12.97% | 38.39% | 32.78% |
| Age at menarche | 12.66 (1.11) | 2.89% | 12.66 | 12.62 (1.17) |
| BMI | 20.21 (3.58) | 21.42% | 20.40 | 20.10 (3.59) |
| SDQ internal | 2.59 (2.60) | 17.12% | 2.63 | 2.71 (2.66) |
| SDQ external | 3.68 (2.91) | 17.09% | 3.74 | 3.77 (2.89) |
| IQ | 104.59 (15.57) | 22.46% | 103.50 | 103.86 (15.98) |

Abbreviations: SD, standard deviation; CSE, certificate of secondary education; BMI, body mass index; SDQ, strengths and difficulties questionnaire; IQ, intelligence quotient.

#### Supplementary Results

**Supplementary Table 3. Linear regression analysis of the association between menstrual symptoms and school absences and GCSE score and logistic regression analysis of the association between menstrual symptoms and achieving five A*-C GCSEs including Maths and English and persistent absence (10% or more) (N=2698).**

| Continuous Outcomes | | | | | | |
|---|---|---|---|---|---|---|
| Exposure | School absence in Year 11 | | | GCSE points score | | |
| | % change | 95% CI | P value | Beta | 95% CI | P Value |
| **Heavy or prolonged bleeding** | | | | | | |
| Crude | 22.71 | 15.20, 30.71 | <0.0001 | -16.97 | -22.70, -11.24 | <0.0001 |
| Adjusted | 16.58 | 9.43, 24.20 | <0.0001 | -5.65 | -10.10, -1.20 | 0.0129 |
| **Menstrual pain** | | | | | | |
| Crude | 16.28 | 9.37, 23.62 | <0.0001 | -3.42 | -8.99, 2.14 | 0.2279 |
| Adjusted | 12.83 | 6.14, 19.95 | 0.0001 | -3.14 | -7.46, 1.17 | 0.1532 |
| **Binary Outcomes** | | | | | | |
| Exposure | Persistent absence in Year 11 | | | Five A*-C GCSEs incl. Maths & English | | |
| | OR | 95% CI | P value | OR | 95% CI | P Value |
| **Heavy or prolonged bleeding** | | | | | | |
| Crude | 1.69 | 1.65, 1.73 | <0.0001 | 0.60 | 0.59, 0.62 | <0.0001 |
| Adjusted | 1.48 | 1.45, 1.52 | <0.0001 | 0.73 | 0.71, 0.75 | <0.0001 |
| **Menstrual pain** | | | | | | |
| Crude | 1.51 | 1.47, 1.54 | <0.0001 | 0.91 | 0.89, 0.92 | <0.0001 |
| Adjusted | 1.42 | 1.39, 1.46 | <0.0001 | 0.84 | 0.81, 0.86 | <0.0001 |

Abbreviations: GCSE, general certificate of secondary education; OR, odds ratio. Adjusted for ethnicity; maternal education, parental social class, financial difficulties, and home ownership during pregnancy; maternal smoking pre-pregnancy; parental separation, physical abuse, and sexual abuse before 11; maternal depression at 12.1; age at menarche; body mass index at 12.8; internalising and externalising problems at 9.6; and intelligence quotient at 8.

**Supplementary Table 4. Linear regression analysis of the association between different categories of menstrual symptoms and GCSE score and school absences (N=2698).**

| | | School absence in Year 11 | | | GCSE points score | | |
|---|---|---|---|---|---|---|---|
| | | % increase | 95% CI | P value | Beta | 95% CI | P Value |
| **Heavy or prolonged bleeding and visiting a doctor** | | | | | | | |
| Crude analysis | No symptom (ref) (N=2353) | 0 | - | - | 0 | - | - |
| | Symptom, no doctor (N=967) | 15.21 | 7.68, 23.27 | <0.0001 | -14.08 | -20.23, -7.93 | <0.0001 |
| | Symptom and doctor (N=246) | 57.36 | 39.53, 77.47 | <0.0001 | -28.29 | -39.22, -17.35 | <0.0001 |
| Adjusted analysis | No symptom (ref) (N=2353) | 0 | - | - | 0 | - | - |
| | Symptom, no doctor (N=967) | 10.93 | 3.70, 18.67 | 0.0026 | -5.45 | -10.23, -0.68 | 0.0252 |
| | Symptom and doctor (N=246) | 42.61 | 26.41, 60.89 | <0.0001 | -5.92 | -14.56, 2.72 | 0.1791 |
| **Menstrual pain and visiting a doctor** | | | | | | | |
| Crude analysis | No symptom (ref) (N=1625) | 0 | - | - | 0 | - | - |
| | Symptom, no doctor (N=1616) | 10.65 | 3.88, 17.86 | 0.0017 | -0.54 | -6.30, 5.23 | 0.8552 |
| | Symptom and doctor (N=292) | 55.64 | 38.70, 74.64 | <0.0001 | -20.08 | -30.6, -9.57 | 0.0002 |
| Adjusted analysis | No symptom (ref) (N=1625) | 0 | - | - | 0 | - | - |
| | Symptom, no doctor (N=1616) | 8.76 | 2.13, 15.83 | 0.0089 | -2.53 | -6.99, 1.94 | 0.2667 |
| | Symptom and doctor (N=292) | 42.30 | 26.74, 59.77 | <0.0001 | -6.77 | -15.00, 1.46 | 0.1068 |
| **Heavy and prolonged, separately and combined** | | | | | | | |
| Crude analysis | Neither (ref) (N=2013) | 0 | - | - | 0 | - | - |
| | Prolonged only (N=286) | -8.48 | -18.37, 2.61 | 0.1290 | 4.78 | -5.59, 15.15 | 0.3663 |
| | Heavy only (N=882) | 20.74 | 12.46, 29.62 | <0.0001 | -17.24 | -23.69, -10.8 | <0.0001 |
| | Both (N=307) | 23.22 | 10.49, 37.42 | 0.0002 | -13.73 | -23.63, -3.84 | 0.0065 |
| Adjusted analysis | Neither (ref) (N=2013) | 0 | - | - | 0 | - | - |
| | Prolonged only (N=286) | -7.54 | -17.39, 3.48 | 0.1723 | 5.15 | -2.84, 13.15 | 0.2064 |
| | Heavy only (N=882) | 14.69 | 6.84, 23.13 | 0.0002 | -4.82 | -9.81, 0.16 | 0.0580 |
| | Both (N=307) | 17.71 | 5.65, 31.14 | 0.0031 | -5.55 | -13.19, 2.10 | 0.1549 |
| **Heavy and pain, separately and combined** | | | | | | | |
| Crude analysis | Neither (ref) (N=1280) | 0 | - | - | 0 | - | - |
| | Pain only (N=952) | 10.44 | 2.31, 19.21 | 0.0109 | 1.18 | -5.76, 8.12 | 0.7388 |
| | Heavy only (N=276) | 19.40 | 6.28, 34.13 | 0.0028 | -19.78 | -30.35, -9.21 | 0.0002 |
| | Both (N=920) | 30.86 | 21.22, 41.26 | <0.0001 | -15.46 | -22.4, -8.51 | <0.0001 |
| Adjusted analysis | Neither (ref) (N=1280) | 0 | - | - | 0 | - | - |
| | Pain only (N=952) | 8.84 | 0.88, 17.42 | 0.0288 | -2.88 | -8.24, 2.48 | 0.2925 |
| | Heavy only (N=276) | 14.21 | 1.78, 28.14 | 0.0238 | -8.18 | -16.34, -0.01 | 0.0498 |
| | Both (N=920) | 23.13 | 14.02, 32.96 | <0.0001 | -6.53 | -11.94, -1.11 | 0.0182 |

Abbreviations: GCSE, general certificate of secondary education. Adjusted for ethnicity; maternal education, parental social class, financial difficulties, and home ownership during pregnancy; maternal smoking pre-pregnancy; parental separation, physical abuse, and sexual abuse before 11; maternal depression at 12.1; age at menarche; body mass index at 12.8; internalising and externalising problems at 9.6; and intelligence quotient at 8.

The following results are from analyses conducted in the complete case samples only.

**Supplementary Table 5. Distribution of sociodemographic factors, adverse childhood experiences, and child factors according to adolescent menstrual symptoms in complete case sample only.**

| | Heavy or Prolonged Bleeding | | Menstrual Pain | |
|---|---|---|---|---|
| | **Yes**<br>≤ 972 (36.03%) | **No**<br>≤ 1726 (63.97%) | **Yes**<br>≤ 1496 (55.45%) | **No**<br>≤ 1202 (44.55%) |
| **SOCIODEMOGRAPHIC FACTORS** | | | | |
| **Ethnicity** | | | | |
| White | 830 (96.51) | 1528 (96.16) | 1301 (96.44) | 1057 (96.09) |
| Non-white | 30 (3.49) | 61 (3.84) | 48 (3.56) | 43 (3.91) |
| **Maternal education** | | | | |
| CSE/Vocational | 195 (23.75) | 306 (19.70) | 276 (21.15) | 225 (21.05) |
| O level | 327 (39.83) | 579 (37.28) | 486 (37.24) | 420 (39.29) |
| A level | 214 (26.07) | 407 (26.21) | 354 (27.13) | 267 (24.98) |
| Degree | 85 (10.35) | 261 (16.81) | 189 (14.48) | 157 (14.69) |
| **Parental social class** | | | | |
| Manual | 423 (50.78) | 681 (44.31) | 609 (46.56) | 495 (46.61) |
| Non-manual | 410 (49.22) | 856 (55.69) | 699 (53.44) | 567 (53.39) |
| **Financial difficulties** | | | | |
| Any | 556 (64.95) | 910 (58.15) | 826 (61.60) | 640 (59.26) |
| None | 300 (35.05) | 655 (41.85) | 515 (38.40) | 440 (40.74) |
| **Home ownership** | | | | |
| Renter or non-homeowner | 144 (16.36) | 192 (12.00) | 194 (14.10) | 142 (12.86) |
| Owner or private renter | 736 (83.64) | 1408 (88.00) | 1182 (85.90) | 962 (87.14) |
| **Maternal smoking pre-pregnancy** | | | | |
| Yes | 277 (30.88) | 400 (24.74) | 409 (29.34) | 268 (23.93) |
| No | 620 (69.12) | 1217 (75.26) | 985 (70.66) | 852 (76.07) |
| **ADVERSE CHILDHOOD EXPERIENCES** | | | | |
| **Parental separation before age 11** | | | | |
| Yes | 212 (25.27) | 329 (21.48) | 300 (22.76) | 241 (22.89) |
| No | 627 (74.73) | 1203 (78.52) | 1018 (77.24) | 812 (77.11) |
| **Physical abuse before age 11** | | | | |
| Any | 317 (38.33) | 593 (38.99) | 533 (40.69) | 377 (36.32) |
| None | 510 (61.67) | 928 (61.01) | 777 (59.31) | 661 (63.68) |
| **Sexual abuse before age 11** | | | | |
| Any | 38 (4.67) | 39 (2.58) | 42 (3.28) | 35 (3.36) |
| None | 775 (95.33) | 1473 (97.42) | 1240 (96.72) | 1008 (96.64) |
| **Maternal depression at 12.1 years** | | | | |
| Yes | 213 (29.42) | 267 (19.31) | 291 (25.15) | 189 (19.89) |
| No | 511 (70.58) | 1116 (80.69) | 866 (74.85) | 761 (80.11) |
| **CHILD FACTORS** | | | | |
| **Age at menarche** | | | | |
| Mean (SE) | 12.60 (1.12) | 12.69 (1.10) | 12.53 (1.08) | 12.83 (1.31) |
| **BMI at 12.8 years** | | | | |
| Mean (SE) | 20.40 (3.54) | 20.11 (3.59) | 20.39 (3.16) | 19.99 (3.64) |
| **Internalising SDQ at 9.6 years** | | | | |
| Mean (SE) | 2.74 (2.72) | 2.51 (2.53) | 2.76 (2.69) | 2.38 (2.47) |
| **Externalising SDQ at 9.6 years** | | | | |
| Mean (SE) | 4.05 (3.08) | 3.49 (2.79) | 3.82 (2.91) | 3.52 (2.91) |
| **IQ at 8 years** | | | | |
| Mean (SE) | 103.31 (15.86) | 105.26 (15.38) | 104.87 (15.36) | 104.23 (15.83) |
| **Past year oral contraception at exposure timepoint** | | | | |
| Yes | 239 (24.95) | 168 (9.85) | 290 (19.58) | 117 (9.89) |
| No | 719 (75.05) | 1538 (90.15) | 1191 (80.42) | 1066 (90.11) |

Abbreviations: SE, standard error; CSE, certificate of secondary education; BMI, body mass index; SDQ, strengths and difficulties questionnaire; IQ, intelligence quotient.

**Supplementary Table 6. Linear regression analysis of the association between menstrual symptoms and school absences and GCSE score and logistic regression analysis of the association between menstrual symptoms and achieving five A*-C GCSEs including Maths and English and persistent absence (10% or more) in the complete case sample only (N=1258).**

| Continuous Outcomes | | | | | | |
|---|---|---|---|---|---|---|
| **Exposure** | **School absence in Year 11** | | | **GCSE points score** | | |
| | % change | 95% CI | P value | Beta | 95% CI | P Value |
| **Heavy or prolonged bleeding** | | | | | | |
| Crude | 14.43 | 4.63, 25.14 | 0.0032 | -13.17 | -20.36, -5.98 | 0.0003 |
| Adjusted | 11.24 | 1.78, 21.57 | 0.0188 | -6.12 | -11.60, -0.64 | 0.0286 |
| **Menstrual pain** | | | | | | |

| | | | | | | |
|---|---|---|---|---|---|---|
| Crude | 15.40 | 6.12, 25.48 | 0.0008 | -2.79 | -9.56, 3.97 | 0.4181 |
| Adjusted | 11.06 | 2.11, 20.79 | 0.0145 | -2.47 | -7.67, 2.72 | 0.3503 |
| **Binary Outcomes** | | | | | | |
| **Exposure** | **Persistent absence in Year 11** | | | **Five A*-C GCSEs incl. Maths & English** | | |
| | OR | 95% CI | P value | OR | 95% CI | P Value |
| **Heavy or prolonged bleeding** | | | | | | |
| Crude | 1.41 | 1.05, 1.90 | 0.0233 | 0.67 | 0.51, 0.88 | 0.0039 |
| Adjusted | 1.27 | 0.93, 1.73 | 0.1290 | 0.81 | 0.57, 1.14 | 0.2184 |
| **Menstrual pain** | | | | | | |
| Crude | 1.48 | 1.10, 1.99 | 0.0087 | 0.94 | 0.72, 1.23 | 0.6671 |
| Adjusted | 1.40 | 1.03, 1.90 | 0.0324 | 0.92 | 0.66, 1.29 | 0.6387 |

Abbreviations: GCSE, general certificate of secondary education; OR, odds ratio. Adjusted for ethnicity; maternal education, parental social class, financial difficulties, and home ownership during pregnancy; maternal smoking pre-pregnancy; parental separation, physical abuse, and sexual abuse before 11; maternal depression at 12.1; age at menarche; body mass index at 12.8; internalising and externalising problems at 9.6; and intelligence quotient at 8.

**Supplementary Table 7. Linear regression analysis of the associations between different categories of menstrual symptoms and school absences and GCSE score in the complete case sample only (N=1258).**

| | | **School absence in Year 11** | | | **GCSE points score** | | |
|---|---|---|---|---|---|---|---|
| | | % increase | 95% CI | P value | Beta | 95% CI | P Value |
| **Heavy doctor** | | | | | | | |
| Crude analysis | No symptom (ref) | 0 | - | - | 0 | - | - |
| | Symptom, no doctor | 8.03 | -1.69, 18.71 | 0.1084 | -10.82 | -18.42, -3.22 | 0.0053 |
| | Symptom and doctor | 52.99 | 25.72, 86.17 | <0.0001 | -24.99 | -40.81, -9.17 | 0.0020 |
| Adjusted analysis | No symptom (ref) | 0 | - | - | 0 | - | - |
| | Symptom, no doctor | 5.92 | -3.54, 16.31 | 0.2277 | -5.84 | -11.63, -0.05 | 0.0482 |
| | Symptom and doctor | 42.22 | 17.03, 72.83 | 0.0004 | -6.34 | -18.41, 5.72 | 0.3023 |
| **Pain doctor** | | | | | | | |
| Crude analysis | No symptom (ref) | 0 | - | - | 0 | - | - |
| | Symptom, no doctor | 11.38 | 2.25, 21.32 | 0.0136 | -0.45 | -7.38, 6.47 | 0.8976 |
| | Symptom and doctor | 55.47 | 29.64, 86.44 | <0.0001 | -22.48 | -37.18, -7.77 | 0.0028 |
| Adjusted analysis | No symptom (ref) | 0 | - | - | 0 | - | - |
| | Symptom, no doctor | 7.97 | -0.89, 17.62 | 0.0793 | -0.97 | -6.27, 4.33 | 0.7194 |
| | Symptom and doctor | 43.28 | 19.38, 71.95 | 0.0001 | -16.05 | -27.34, -4.77 | 0.0053 |
| **Heavy and prolonged, separately and combined** | | | | | | | |
| Crude analysis | Neither (ref) | 0 | - | - | 0 | - | - |
| | Prolonged only | -9.63 | -22.36, 5.18 | 0.1906 | 1.77 | -10.43, 13.96 | 0.7761 |
| | Heavy only | 12.58 | 1.87, 24.43 | 0.0203 | -14.41 | -22.45, -6.37 | 0.0005 |
| | Both | 14.28 | -2.95, 34.57 | 0.1093 | -8.04 | -21.17, 5.10 | 0.2301 |
| Adjusted analysis | Neither (ref) | 0 | - | - | 0 | - | - |
| | Prolonged only | -8.82 | -21.54, 5.96 | 0.2282 | 5.40 | -3.87, 14.67 | 0.2530 |
| | Heavy only | 8.79 | -1.49, 20.14 | 0.0962 | -6.38 | -12.50, -0.26 | 0.0412 |
| | Both | 13.79 | -3.21, 33.78 | 0.1174 | -2.24 | -12.22, 7.74 | 0.6603 |
| **Heavy and pain, separately and combined** | | | | | | | |
| Crude analysis | Neither (ref) | 0 | - | - | 0 | - | - |
| | Pain only | 10.99 | 0.32, 22.79 | 0.0433 | 3.59 | -4.55, 11.72 | 0.3875 |
| | Heavy only | 6.80 | -9.56, 26.11 | 0.4378 | -5.92 | -19.30, 7.46 | 0.3854 |
| | Both | 24.44 | 11.63, 38.72 | 0.0001 | -13.28 | -22.02, -4.53 | 0.0030 |
| Adjusted analysis | Neither (ref) | 0 | - | - | 0 | - | - |
| | Pain only | 7.10 | -3.20, 18.49 | 0.1835 | 0.74 | -5.51, 6.98 | 0.8172 |
| | Heavy only | 4.38 | -11.42, 23.00 | 0.6083 | -1.44 | -11.58, 8.70 | 0.7807 |
| | Both | 18.32 | 6.14, 31.90 | 0.0024 | -7.16 | -13.87, -0.46 | 0.0364 |

Abbreviations: GCSE, general certificate of secondary education. Adjusted for ethnicity; maternal education, parental social class, financial difficulties, and home ownership during pregnancy; maternal smoking pre-pregnancy; parental separation, physical abuse, and sexual abuse before 11; maternal depression at 12.1; age at menarche; body mass index at 12.8; internalising and externalising problems at 9.6; and intelligence quotient at 8.
